# DNA methylation patterns contribute to changes of cellular differentiation pathways in leukocytes with LOY from patients with Alzheimer’s disease

**DOI:** 10.1101/2024.08.19.24312211

**Authors:** Marcin Jąkalski, Bożena Bruhn-Olszewska, Edyta Rychlicka-Buniowska, Hanna Davies, Daniil Sarkisyan, Maciej Siedlar, Jarosław Baran, Kazimierz Węglarczyk, Janusz Jaszczynski, Janusz Ryś, Vilmantas Gedraitis, Natalia Filipowicz, Alicja Klich-Rączka, Lena Kilander, Martin Ingelsson, Jan P. Dumanski

## Abstract

Alzheimer’s disease (AD) is a common and increasing societal problem due to the extending human lifespan. In males, loss of chromosome Y (LOY) in leukocytes is strongly associated with AD. We studied here DNA methylation and RNA expression in sorted monocytes and granulocytes with and without LOY from male AD patients. Through multi-omic analysis, we identified new candidate genes and confirmed the involvement of numerous genes previously associated with AD. Our findings highlight LOY-related differences in DNA methylation that occur in gene regulatory regions and are predominantly accompanied by down-regulation of affected genes. Specifically, we observed alterations in key genes involved in leukocyte differentiation: *FLI1*, involved in early hematopoiesis; *RUNX1*, essential for blood cell development; *RARA*, regulating gene expression in response to retinoic acid; *CANX*, crucial for protein folding; *CEBPB*, a transcription factor important for immune responses; and *MYADM*, implicated in cell adhesion and migration. Moreover, protein-protein interaction analysis in granulocytes identified that products of two of these genes, *CANX* and *CEBPB*, are key hub proteins. Thus, LOY appears to dysregulate genes involved in leukocyte differentiation and induce higher-level epigenetic changes. This research underscores the potential of multi-omic approaches in pure cell populations to uncover the molecular underpinnings of AD and reinforces the significance of LOY as a pathogenic factor in this disease. Overall, results support the hypothesis that age-related immune cell dysfunction contributes to AD development. Finally, our results link previous analysis showing impact of LOY on leukocyte differentiation, LOY-associated transcriptional dysregulation and GWAS studies of LOY.

## INTRODUCTION

Loss of chromosome Y (LOY) in leukocytes from aging males [1] is the most common post-zygotic mutation, detectable in whole blood DNA from >40% of men above the age of 70 years [2], reaching 57% in the analysis of bulk DNA from 93-year-old men [3]. A single-cell analysis of Peripheral Blood Mononuclear Cells (PBMCs) derived from 29 aging men (median age 80 years) identified cells with LOY in every studied subject [4]. The presence of LOY has also been reported in other tissues although with lower frequencies [3, 5]. Importantly, a serial analysis of blood samples showed that LOY is a dynamic process [1, 6, 7].

Major risk factors for LOY include age, smoking, and germline predisposition as well as environmental and occupational hazards [1, 2, 8–14]. It has also been suggested that LOY affects different lineages of hematopoietic cells with different frequencies and that it plays a role in the dysregulation of autosomal genes through LOY-Associated Transcriptional Effects (LATE) in a pleiotropic manner [4]. Moreover, dysregulation of large sets of various immune genes was pronounced in LOY cells [4, 15–17].

LOY has been associated with increased risk for all-cause mortality as well as with chronic and acute age-related diseases inside and outside of the hematopoietic system, with causal effects of LOY already shown for cardiac fibrosis and bladder cancer [1, 5, 7, 18–21]. Notably, a strong association between LOY and late-onset Alzheimer’s disease (LOAD) was observed, as males with LOY had a 6.8-fold greater risk for Alzheimer’s disease (AD) diagnosis [9]. This effect is comparable to that of the strongest genetic risk factor for LOAD, the *ε*4 allele of the apolipoprotein E gene (*APOE*). The presence of one or two copies of *APOE ε*4 increases the risk of developing LOAD by a factor of 3 up to 15-fold in a dose-dependent manner [22, 23]. The replication of the association between LOY and AD in additional cohorts, also applying a different methodological approach, has recently been shown in two reports [24, 25]. Transcriptome analyses further suggested that LOY might indeed play an important role in the pathogenesis of AD [15, 26]. Specifically, a recent study of LOY in human brain tissues from healthy aging subjects has shown that microglia have the highest percentage of LOY and remarkably, there was a significant increase of LOY in microglia from male AD donors[15]. Brain microglia and circulating monocytes represent functionally closely related cells, and monocytes from blood can migrate across the blood-brain barrier (BBB) in response to inflammatory stimuli in various diseases, including AD [27–30]. The process of clearance of amyloid plaques by microglia in the brain and clearance of circulating amyloid-beta from blood by monocytes has further been suggested as an important disease mechanism in AD [31, 32]. Moreover, apart from the impact of monocytes in AD pathogenesis, neutrophils, the most abundant granulocyte cells, exhibit a hyperactive phenotype by elevated production of reactive oxygen species (ROS) in AD, leading to inflammation and disease progression [33].

Alzheimer disease is the most common neurodegenerative disorder and a major public health problem, which is expected to worsen due to the ever-increasing lifespan [34]. Depending on whether the onset of symptoms occurs before or after 65 years of age, AD can be broadly defined into LOAD (∼90% of AD patients) and early onset AD (EOAD)[22]. Whereas EOAD sometimes can be explained by mutations in either of three genes (*APP, PSEN1, PSEN2*), LOAD is believed to be caused by complex genetics in combination with environmental factors [35]. For LOAD, analyses of twins have suggested a high heritability of AD with estimates ranging between 60-80% [36]. In addition to *APOE,* genome-wide association studies (GWAS) have identified several other AD risk genes [22, 35, 37, 38].

Multiple studies have also shown that epigenetic processes are often dysregulated and might play a role in the development and progression of AD. DNA methylation at CpG sites provides a stable epigenetic modification that usually silences (alternatively promotes in case of demethylation) the transcription of adjacent gene(s) [39, 40]. Altered DNA methylation levels in whole blood DNA were identified as associated with worse cognitive performance and accelerated rate of AD progression [41–44]. Furthermore, covalent modifications of histones, and higher-level three-dimensional structures (such as topologically associating domains, TADs), are other examples of dynamic epigenetic regulators. TADs constitute one of the forms of chromosomal organization within a nucleus into functional compartments [45]. Genes bound by the same TAD are usually regulated in a coordinated manner, as TADs facilitate interactions between the genes and their distant regulatory elements. DNA methylation and histone modifications, such as histone methylation and acetylation, are in a constant interplay [46, 47]. Recent studies showed that epigenetic processes are often dysregulated in AD patients and that the epigenetic control of enhancers may have an important pathogenetic role [47–49].

We hypothesized that LOY may be associated with global changes in DNA methylation and may lead to the discovery of new, specific candidate genes. Therefore, we have here taken a novel approach to study AD by incorporating multi-omics data (DNA methylation and gene expression) and the LOY status of the studied individuals, to delineate candidate genes that might be involved in the pathogenesis of this condition. For this purpose, we took advantage of pure flow cytometry-sorted populations of granulocytes and monocytes (with at least 95% purity) with or without LOY.

## MATERIALS AND METHODS

### Study group

Blood samples from patients diagnosed with AD were collected from male subjects in Uppsala, Sweden and Kraków, Poland for the purpose of estimating LOY levels as described [4]. The availability of sufficient amount of DNA was used to select a group of 43 individuals for complementing the previous study with DNA methylation analyses. Samples from AD patients were collected during January 2015 to May 2018, at the Geriatric/Memory Clinic, Uppsala Academic Hospital Sweden. Additional samples from AD patients were collected from January 2017 to May 2018 at the Clinic of Internal Diseases and Gerontology of the Jagiellonian University in Kraków. The criteria for recruitment of AD patients were ongoing clinically and radiologically confirmed diagnosis, intermediate or severely advanced disease.

The study was approved by the local research ethics committee in Uppsala, Sweden (Regionala Etikprövningsnämnden i Uppsala (EPN): Dnr 2005-244, Ö48-2005; Dnr 2013/350; Dnr 2015/092; Dnr 2015/458; Dnr 2015/458/2, the latter with update from 2018) and the Bioethical Committee of the Regional Medical Chamber in Kraków, Poland (No. 6/KBL/OIL/2014). All participants or next of kin have given their written informed consent to participate. The study was conducted according to the guidelines of the Declaration of Helsinki.

### Sample preparation

16 ml of blood was collected into two BD Vacutainer® CPT™ Mononuclear Cell Preparation Tubes (BD). Peripheral blood mononuclear cells (PBMCs) were isolated following the manufacturer’s instructions. The PBMCs were then washed with PBS, a portion of PBMCs were aliquoted for scRNA-seq. Additional 16 ml of whole blood were collected into two BD Vacutainer® K2 EDTA tubes (BD). Red blood cells were lysed using 1× BD Pharm Lyse™ lysing solution (BD). Isolated white blood cells (WBCs) were washed with PBS. Targeted cell populations were sorted from the isolated PBMCs and WBCs using FACS [4]. In brief, live cells were sorted based on their FSC and SSC. Monocytes were defined based on their size and as CD14+; granulocytes were defined based on their size and granularity. Cells were sorted to achieve the purity of above 96%. Cell fractions were split for downstream DNA or RNA extractions. Cells sorted for RNA extraction was dissolved in RNAprotect Cell Reagent (Qiagen), all cell fractions were then pelleted and frozen in −70 for further processing.

### Estimation of LOY levels

DNA was extracted and quantified from each isolated cell population following established protocol [4]. DNA was genotyped using three different versions of SNP-arrays InfiniumCoreExome-24v1-1, InfiniumOmniExpressExome-8v1-3 and InfiniumQCArray-24v1 (Illumina). All genotyping experiments were performed following the manufacturer’s instructions at the Science for Life technology platform SNP&SEQ at Uppsala University, Sweden. All included experiments passed strict quality control at the genotyping facility. Additional QC criteria as well as calculation of mLRRY were done as described [4]. The percentage of cells with LOY (%LOY) in each sample was estimated using the published formula, i.e., 100*(1-(2^(2*mLRRY)^)) [6].

### Bisulfite conversion of DNA samples

Based on the DNA concentrations determined by Quant-iT PicoGreen dsDNA Assay (Thermo Scientific), 250 ng of each DNA sample were used in bisulfite conversion of methylated CpG sites using the EZ DNA MethylationTM Kit (Zymo Research). The bisulfite converted DNA was eluted used for methylation analysis according to the manufacturer’s protocol.

### Methylation analysis

Methylation profiling was performed with the Infinium assay using the MethylationEPIC_v-1-0 array (Illumina) according to the manufacturer’s protocol. The scanning of the EPIC arrays and determination of signal intensities were performed by the iScan System (Illumina). Intensities were normalized using Illumina’s internal normalization probes and algorithms, with background subtraction.

### Analysis of DNA methylation data

The raw IDAT files were read into R using the readEPIC function from the wateRmelon package. Next, lumiMetyC (with quantile normalization) and BMIQ functions from the lumi and wateRmelon packages respectively were used to perform data normalization. The data was filtered by minimum detection p-value and all probes overlapping with known SNPs were removed. Probe annotation was performed within R using the package *IlluminaHumanMethylationEPICanno.ilm10b4.hg19*. Here we focused on several different annotation categories, such as *UCSC_RefGene_NAME, Relation_to_Island* (OpenSea, Island, N_Shore, N_Shelf, S_Shore, S_Shelf), *Regulatory_Feature_Group* (Promoter_Associated, Gene_Associated, NonGene_Associated, Unclassified), *UCSC_RefGene_Group* (TSS1500, TSS200, 5’UTR, 1stExon, Body, ExonBnd, 3’UTR).

To identify statistically significant differences in methylation between the compared groups we used the *dmpFinder* function from the *minfi* package. Probes located on chromosome Y were removed prior to the analysis. Significant DMPs were called when absolute average change in methylation between sample groups was at least 0.5 (M-value) and adjusted P-value<0.05. To intersect DMPs with gene regions, we used the UCSC based annotations and the promoter regions of UCSC genes were defined by a genomic window of +/- 2 kb from TSS using the *promoters* function from the GenomicRanges R package [50]. Visualization of genomic neighborhood was performed using *Gviz* package. Gene annotations were plotted for *hg19* version of the human genome using UCSC-based gene annotations.

### Bulk RNA extraction and sequencing

RNA-seq data of sorted cell populations were processed as described previously [4]. Briefly, cell pellets were used for extraction of RNA using RiboPure™ RNA Purification Kit (Thermo Fisher Scientific), which was further purified from possible DNA contaminations with TURBO DNA-free™ Kit (Thermo Fisher Scientific), and finally followed by quality assessment with Agilent 2100 Bioanalyzer (Agilent Technologies). Libraries of all samples were prepared using the Ion AmpliSeq Human Gene Expression kit (Thermo Fisher Man0010742), loaded on Ion 550 chips, and sequenced on Ion S5 XL system. All sequencing library handling steps were done using manufacturers’ recommendations.

### Bulk RNA-seq data processing

The sequenced AmpliSeq data were basecalled with the Ion Torrent Suite Sever 5.8.0.RC2 software (Thermo Fisher). The generated reads were aligned to the human transcriptome reference (hg19 AmpliSeq Transcriptome ERCC v1) with TMAP mapper. Next, the raw gene expression counts for all amplicon targets in the assay (20,183) were merged to create separate count matrices for each of the two cell types (granulocytes and monocytes). Count data were processed with the R library *edgeR* version 3.28.1 [51]. We kept only the genes with expression levels above 1 count per million in at least six samples to remove low quality data. Further assessment of data quality using principal component plots revealed sample grouping corresponding to sequencing batch and patient source. The batch effects were thus adjusted using the *ComBat_seq* function from the *sva* package (version 3.35.2) [52]. Genewise Negative Binomial Generalized Linear model was applied to test for differential expression of genes between the AD patients with and without LOY. We considered genes to be significantly differentially expressed by applying a threshold of <0.05 to the corrected p-values (FDR, Benjamini-Hochberg adjustment).

### Annotation of genes, gene ontologies and KEGG pathways

Gene set enrichment analyses were performed in R using the *Cluster Profiler* package. Gene symbols (either DMG or DEG) were mapped to Entrez ID. As a background for enrichment analyses for DEGs we used a set of all genes expressed in each tissue (based on bulk RNA-seq data). DMGs were tested against the background of all genes.

### Analyses of protein-protein interaction networks using STRING

We used the STRING database to identify if any protein-protein interaction network existed among the genes identified in our study. Here we decided to utilize only genes showing alterations of DNA methylation within their promoter region and following the canonical model of DNA methylation vs. expression change. Thus, we applied 157 and 10 genes from granulocytes and monocytes, respectively, to the search box “multiple proteins” in the STRING database server (https://string-db.org/). Disconnected nodes, as well as nodes with less than 3 connections were removed [53].

### Analyses of transcription factor binding site enrichment

We used SEA (Simple Enrichment Analysis) from the MEME package to identify motifs that are relatively enriched among the promoter regions (+/- 2kb from transcription start site) of genes that are both differentially methylated and differentially expressed [54]. Search for enriched motifs was performed against promoter regions of non-differentially methylated genes using the HOCOMOCO database of transcription factor binding motifs (version 11, core HUMAN collection, 401 motifs) [55]. Due to the limitations of the MEME online server, the analyses were run locally using the stand-alone version of the MEME suite (version 5.5.5).

## RESULTS

### Study subjects and measurements of LOY

In order to investigate the effect of LOY on CpG methylation in the context of AD, we analyzed blood samples from 43 men with LOAD (median age 78 years, age range 63-90 years). The leukocytes were sorted using fluorescence-activated cell sorting (FACS) to obtain pure cell populations of granulocytes and monocytes. The samples studied were selected from previously reported individuals [4] and the only selection criterion was the availability of a sufficient amount of DNA, to perform CpG methylation profiling (Figure 1). The status of LOY mosaicism for each cell type was assessed using SNP arrays, following a previously described method [1, 9]. The obtained mLRRY values (median Log R Ratio values of probes located in the male-specific part of chromosome Y) were transformed to estimate the percentage of LOY (%LOY) in each sample [6]. In total, LOY status was determined for 39 and 24 samples of granulocytes and monocytes, respectively (Figure 2). We used 30% cutoff (*i.e.,* 30% of the cells having LOY, as presented previously [6, 9] to divide the samples into “AD-LOY” and “AD-ROY” (the latter standing for **<u>R</u>**etention **<u>O</u>**f chromosome **<u>Y</u>**) groups (Supplementary File 1 - Table S1). This revealed two separate clusters of data (Figure 2B). Matched samples from granulocytes and monocytes (collected from the same patients) presented a high concordance of the %LOY estimates (Pearson correlation = 0.97, Figure 2B).

**Figure 1.**
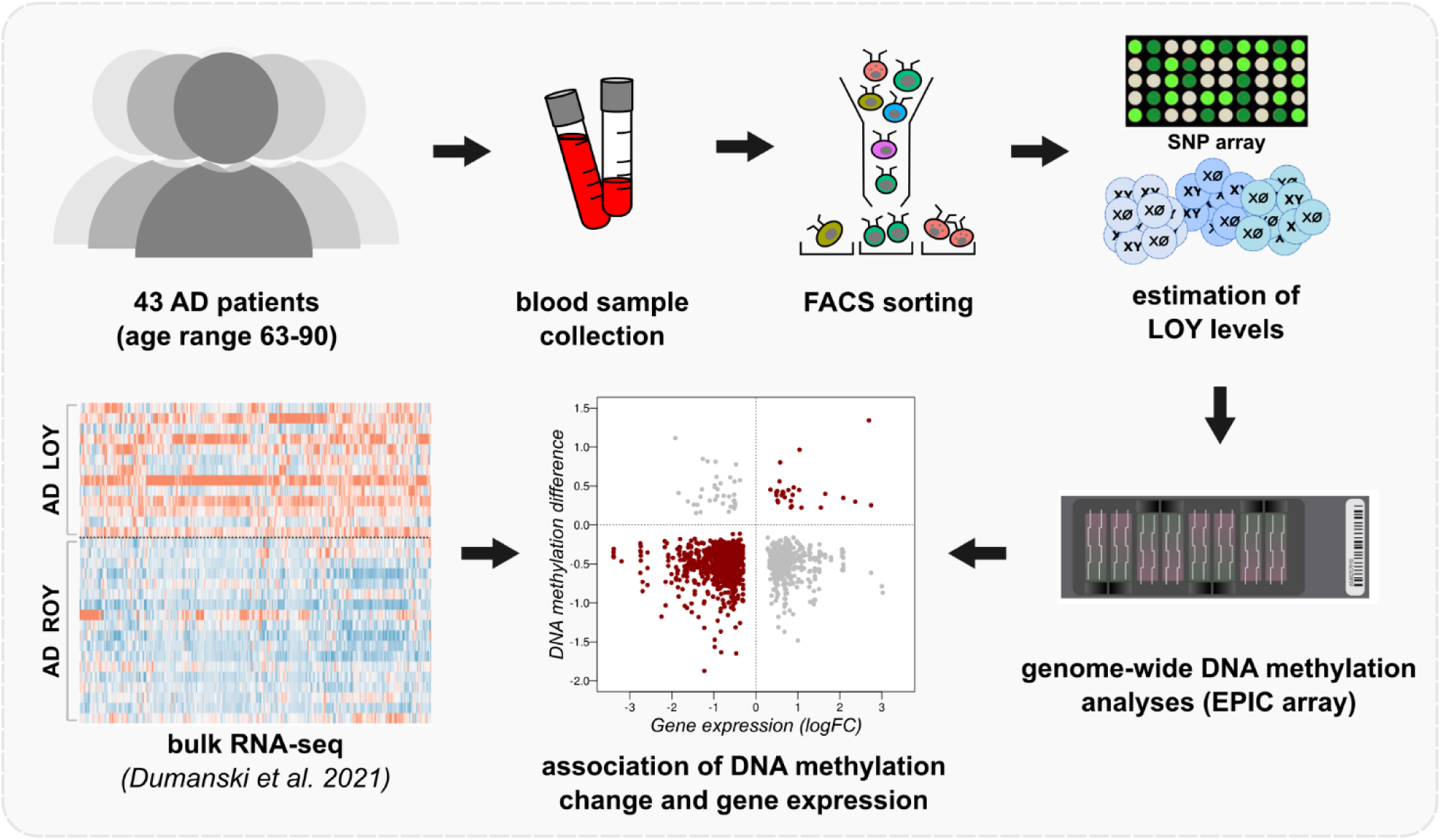
Graphical abstract of the study. We selected 43 AD patients and collected their blood. Leukocytes were then sorted on FACS for two main groups of cells, i.e. granulocytes and monocytes. Estimation of %LOY in each sample (% of cells without Y chromosome) was done using SNP array. ROY stands for **<u>R</u>**etention **<u>O</u>**f chromosome **<u>Y</u>** group of samples. In parallel, samples were subjected to genome-wide DNA methylation profiling using the Illumina EPIC Beadchip. We further incorporated bulk RNA-seq data from our previous project for correlation of variation in DNA methylation with the gene expression levels in samples from the same patients.

**Figure 2.**
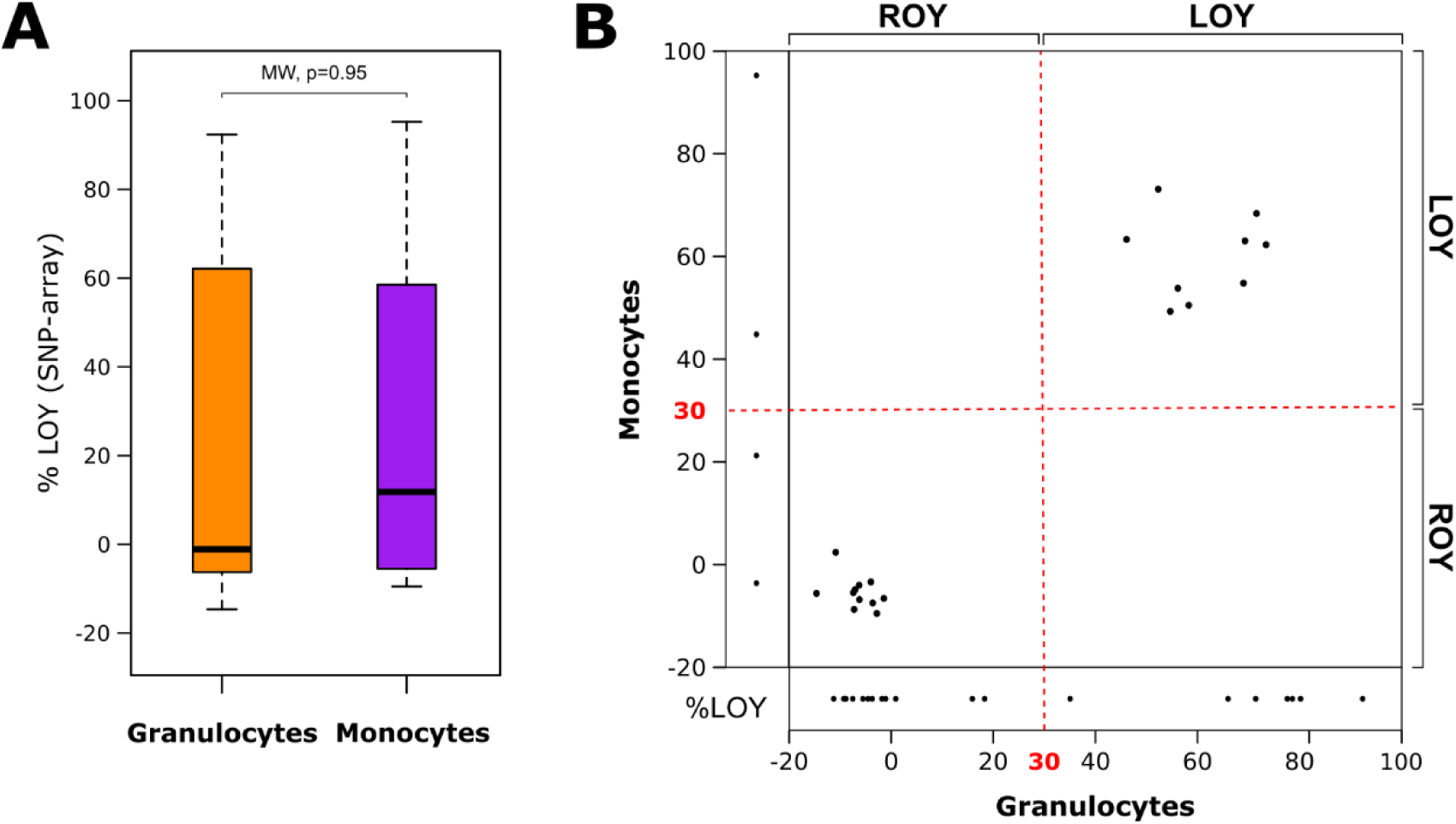
Distribution of LOY levels among the studied samples. (**A**) General distribution of %LOY cells among the two sampled cell types. P-value from Mann–Whitney (MW) pairwise test is present at the top, which not significant, no major differences in %LOY between granulocytes and monocytes. (**B**) Distribution of %LOY in granulocytes (X-axis) and monocytes (Y-axis). Every point in the main panel represents a measurement of %LOY for matched samples collected from the same patients. Narrow panels along each of the axes show results for LOY levels for unmatched samples in each cell type, due to insufficient DNA availability. Red dotted line denotes a 30% threshold used to divide the samples into LOY and ROY groups.

### Genome-wide profiling of DNA methylation landscape

Using the methylation EPIC Beadchip we performed genome-wide DNA methylation analyses among the AD patients. Specifically, we obtained methylation data for 39 granulocyte and 24 monocyte samples covering 684,663 CpG sites, after dropping the problematic probes with known SNPs [56] and those related to smoking [57]. To identify LOY-related DNA methylation changes we performed a differential methylation analysis by comparing AD-LOY against AD-ROY groups. CpG probes located on the Y chromosome were also excluded from the analyses (see Methods). In granulocytes and monocytes we respectively identified 15,269 and 340 significantly differentially methylated probes (hereafter DMPs; adjusted p-value <0.05 and absolute M-value difference >0.1, Table 1, Supplementary File 1 – Tables S2 and S3). As shown in Figure 3A, hypomethylation in AD-LOY samples dominated, both in granulocytes (13,994 of 15,269 DMPs, 92%) and monocytes (309 of 340 DMPs, 91%). Of note, 258 DMPs were shared between granulocytes and monocytes by comparing AD-LOY to AD-ROY samples among AD patients (Figure 3B). Investigation of the genomic distribution of the identified DMPs showed that they were mainly localized within the gene body, intergenic regions, and the so-called open sea regions (see Figure S1).

**Figure 3.**
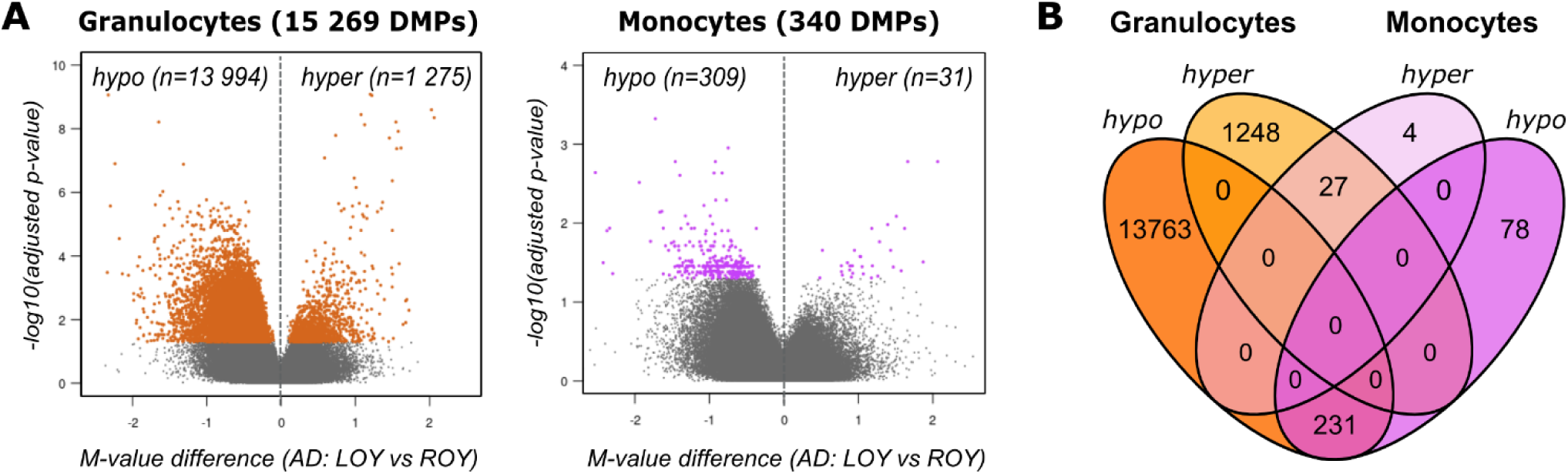
Results from analyses of differential methylation in LOY vs. ROY among AD patients. (**A**) Volcano plots of the genome-wide differential methylation analyses in granulocytes and monocytes. Black points denote all tested CpG sites. Colored dots represent the significant DMPs (FDR <0.05, avDiff >0.1). The direction of methylation change (X-axis) is shown in reference to the LOY samples (hypo – less methylated in cells with LOY, hyper – more methylated in cells with LOY). (**B**) Venn diagram comparing the number of hyper- and hypomethylated DMPs in granulocytes and monocytes identified by comparing LOY to ROY samples among AD patients.

### Gene-associated methylation changes

We next analyzed genes associated with the identified DMPs, *i.e.* differentially methylated genes (DMGs) between AD-LOY and AD-ROY groups (Supplementary Figure S2A and B, Supplementary File 1 – Tables S8-S9). In total, we found 7,105 DMGs in granulocytes. Importantly, 4,177 of these genes had at least one DMP within their promoters, defined as +/- 2 kb from transcription start site, (TSS). The corresponding numbers for monocytes were 252 and 142 DMGs. Granulocytes and monocytes shared a total of 228 DMGs (Supplementary Figure S2C).

Numerous DMGs (∼43-45% in granulocytes and ∼45-49% in monocytes) were associated with AD according to the OpenTargets [58] and GeneCards [59] databases, respecively (Supplementary File 1, Table S9 and S10, Supplementary Figure S3). We conducted a hypergeometric test and found that AD-associated genes were significantly enriched among DMGs, both using OpenTargets (granulocytes: p-value < 2.2e-16; monocytes: p-value=3.0e-10) and GeneCards (granulocytes: p-value=5.2e-05; monocytes: p-value=0.019) databases of AD genes. Additionally, 179 DMGs in granulocytes and 8 DMGs in monocytes were previously found to exhibit LOY-associated dysregulation (Supplementary Figure S3) [4].

### CpG methylation status is linked with changes in gene expression

To further explore the effects of DNA methylation changes, we performed a transcriptomic analysis using bulk RNA sequencing (RNA-seq). The analysis was conducted on available RNA from granulocyte and monocyte samples (31 individuals, see methods). We identified 1,953 and 3,097 genes demonstrating significant differential expression (DEGs) in granulocytes and monocytes, respectively (Figure 4A, Table 7, Supplementary File 1 – Table S4-S5). While our differential methylation analyses showed that hypomethylated DMPs dominated, here the LOY-associated effect on gene expression in AD was less pronounced.

**Figure 4.**
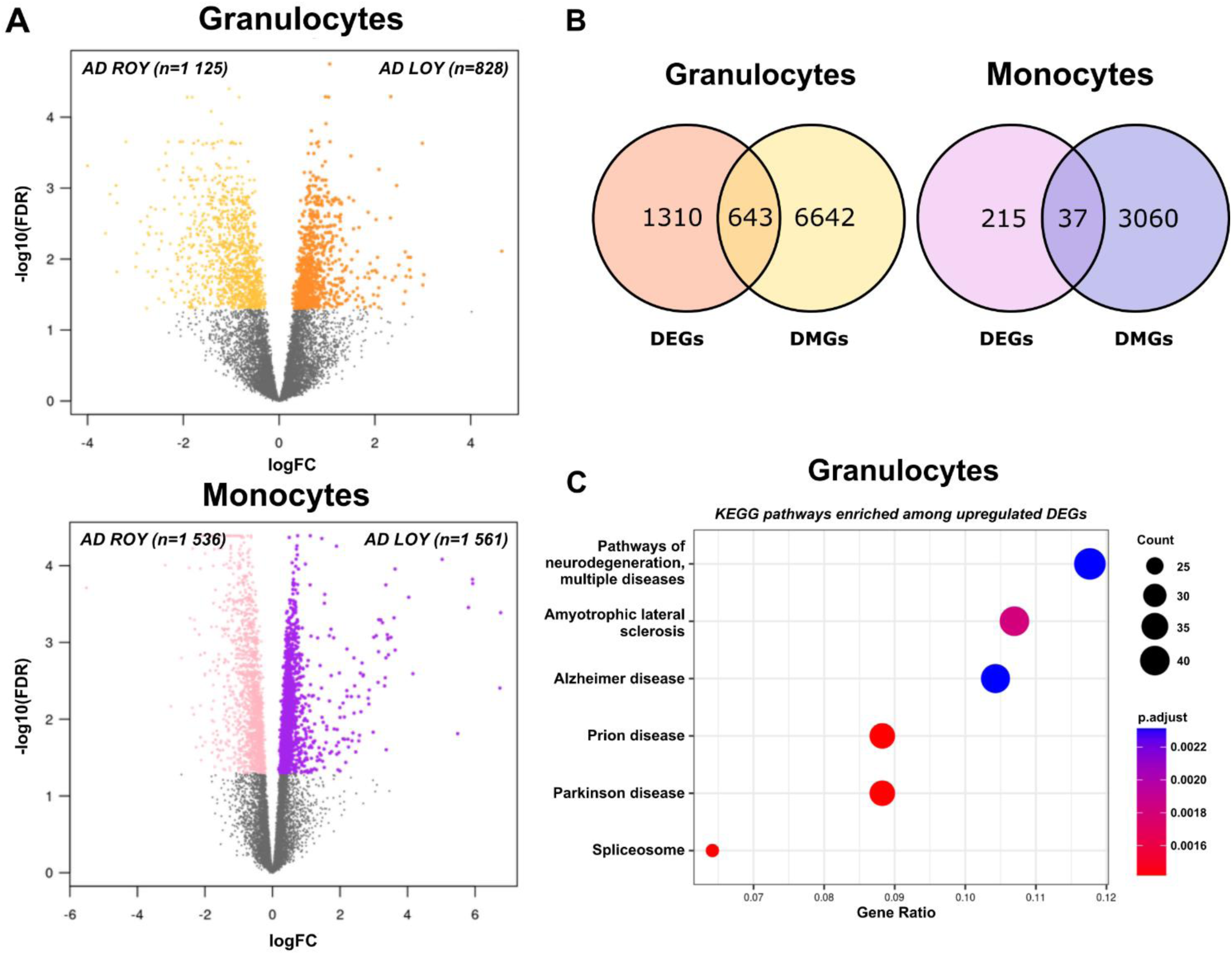
Results from differential expression analyses and the correspondence of promoter-related methylation with the observed expression patterns. (**A**) Volcano plots of the genome-wide differential expression analyses using bulk RNA-seq data from corresponding patients’ samples. Colored dots represent genes with significant change in expression (DEGs, FDR <0.05). Direction of expression change (X-axis) refers to the LOY samples (up – higher in LOY, down – lower in LOY). (**B**) Venn diagrams comparing gene sets derived from the differential expression and differential methylation analyses. (**C**) Results of KEGG pathway enrichment analyses for DEGs upregulated in AD-LOY granulocytes. Size of the circle corresponds to the number of genes representing the given pathway. Color of the circle denotes the adjusted p-value from the enrichment analysis.

We further assessed to what extent DMGs and DEGs overlapped. In granulocytes, 643 genes were shared between the two analyses, and 223 of these had at least one LOY-associated DMP located within the promoter (Figure 4B, Supplementary File 1 – Table S6). The corresponding numbers for monocytes were 37 and 13 (Figure 4B, Supplementary File 1, Table S7). Among granulocytes, DNA methylation of 380 (out of 545, 70%) probes were negatively correlated with the expression level of the associated gene (Supplementary File 1 – Table S8), which is in line with the canonical model, where hypomethylation within CpG islands and promoter region is associated with increased gene expression [39]. Similarly, 13 DEGs in monocytes had differentially methylated sites within their promoters, and methylation changes of 11 of 15 probes (73%) were negatively correlated with the expression of the associated gene (Supplementary File 1 – Table S8). We tested whether overlapping sets of DMGs and DEGs were enriched in any metabolic pathways. We found that genes upregulated in granulocytes were indeed enriched in KEGG pathways associated with neurodegeneration, Parkinson’s disease, and AD (Figure 4C). Detailed results of pathway enrichment analysis are shown in Supplementary File 1, Table S14.

### Examples of DMGs with LOY-associated transcriptional effect

We show selected, representative examples of genes from granulocyte analysis that follow the canonical model of methylation (Figure 5). Additionally, according to the GeneCards database, all of these genes are highly related to AD. One of the genes meeting the canonical model is *CEBPB* (CCAAT enhancer binding protein beta), the expression of which was significantly upregulated in the AD-LOY group in granulocytes (logFC=1.55, FDR=0.043). This one-exon gene harboring twelve CpG probes, had two statistically significant hypomethylated sites identified, both located upstream of its TSS (M-value difference 0.44-0.52, FDR=0.008-0.043) (Figure 5A, 5B). Notably, C/EBP-β is an essential transcription factor during emergency granulopoiesis, and its level has been reported to be elevated in AD brain tissue compared to non-demented control subjects [60, 61]. Moreover, it has been shown that C/EBPβ is upregulated in hematopoietic stem/progenitor cells (HSPCs) under stress conditions in a mouse model [62]. Analogously, the *CANX* (Calnexin) gene illustrates the same pattern of DNA methylation and its gene expression in granulocytes. Our analysis showed that *CANX* was significantly upregulated in LOY granulocytes (logFC=0.91, FDR=0.008) and had a single hypomethylated site located within the promoter region (M-value difference 0.42, FDR=0.023) (Figure 5A, 5B). Calnexin is involved in protein folding, functioning as a chaperone in the endoplasmic reticulum (ER) and this gene is upregulated in the frontal cortex of AD patients [63].

**Figure 5.**
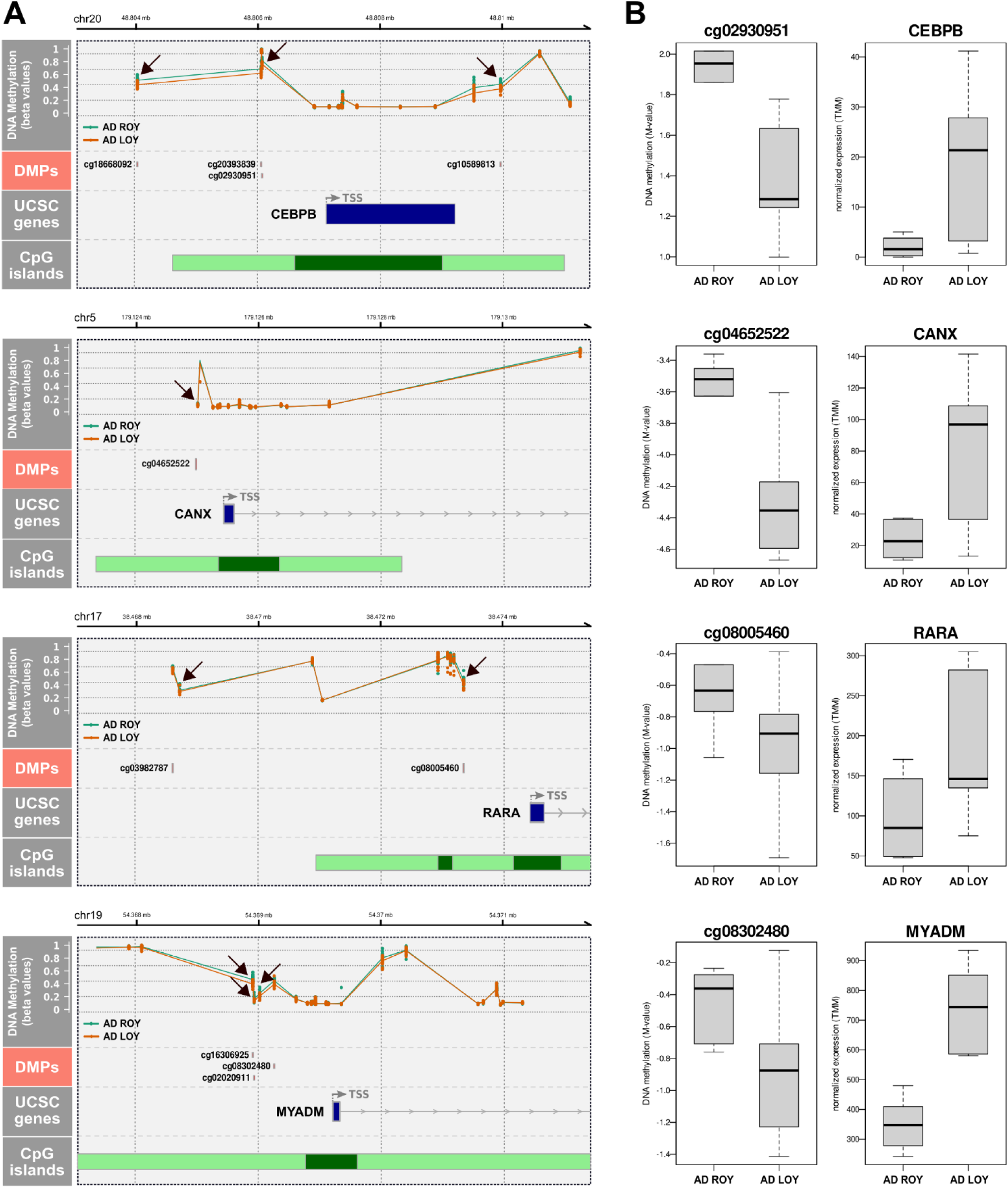
Genomic neighborhood of selected genes from granulocytes with changes in both DNA methylation and gene expression. **(A)** Genomic neighborhood of the promoter region of selected differentially expressed genes from granulocytes. All genes are upregulated in the LOY group in AD patients and have a hypomethylated site within their promoters. Top panel (DNA methylation) shows neighboring CpG probes located in the vicinity of the promoter region and their DNA methylation values in LOY (orange) and ROY (green) samples in AD. Black arrows point to differentially methylated probes. Second panel from top shows the identified significant differentially methylated sites. Panel named “UCSC genes” shows annotations of the gene’s structure. Blue rectangles show exons and introns are shown as thin lines. Arrows on introns show the direction of the gene from its 5’ side. TSS denotes the site of the start of transcription. The bottom-most panel shows CpG islands located in the vicinity of TSS. Dark green denotes CpG islands, and their shores are in light green. Chromosome name and the genomic coordinates are shown above each plot. **(B)** Promoter-associated CpGs with significant change in methylation in granulocytes juxtaposed with a relevant change in expression of the associated gene. Boxplots show methylation and expression values in one of the two compared groups – LOY or ROY in AD patients. Hypomethylation in LOY is associated with upregulation of the corresponding gene’s expression in LOY.

Another interesting example is the *RARA* gene (Retinoic Acid Receptor Alpha), which was upregulated in LOY cells in granulocytes (logFC=0.74, FDR=0.042) and contained a single promoter-related DMP (M-value difference 0.54, FDR=0.035). Additionally, we detected four other DMPs localized outside of the promotor region of this gene, which were all hypomethylated as well (Figure 5A, 5B and Supplementary File 1, Table S2 and S6). A decline in the transcriptomic levels of retinoic acid receptors was suggested to be involved in the early stages of AD mouse models [64]. For the myeloid-associated differentiation marker gene *MYADM*, our analysis showed that three hypomethylated probes were identified in the promotor region of this gene (M-value difference 0.37-0.50, FDR=0.121-0.037). Also, *MYADM* was upregulated in LOY-harboring granulocytes (logFC=0.622, FDR=0.0153) (Figure 5A, 5B). This gene has been reported as upregulated during myeloid differentiation [65].

### Genes with LOY-associated changes in DNA methylation and expression are part of a large interaction network

In order to characterize the interaction between genes that exhibited variability in both DNA methylation and gene expression, we performed protein-protein interaction (PPI) analysis using the STRING database (see Methods). The resulting PPI network in granulocytes consisted of 105 edges and 153 nodes, representing 79 known and 7 predicted protein-protein interactions (Figure 6A, Supplementary File 1, Table S11). We found two hub proteins with the highest number of connections encoded by the *CANX* and *CEBPB* genes, as the representatives of two important interaction networks (Figure 6B and 6C, respectively). Specifically, the *CANX* (calnexin, calcium-binding protein) gene showed 12 known interactions with other products of DMG/DEG genes from granulocytes, while the *CEBPB* (CCAAT enhancer binding protein beta) gene had 11 known interactions with other products of DMG/DEG genes from granulocytes. No network could be identified for the gene set from monocytes.

**Figure 6.**
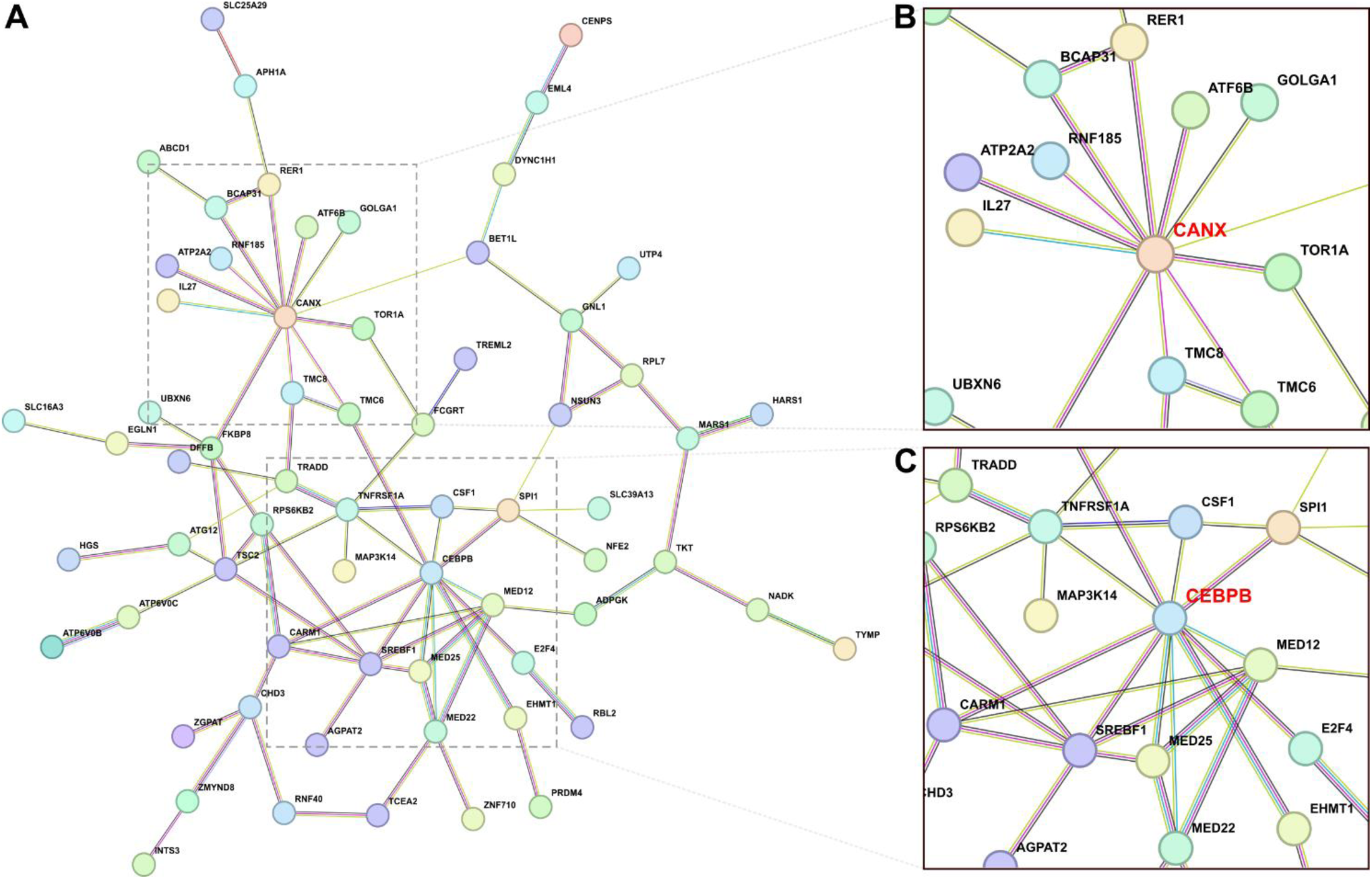
String protein-protein interaction network based on genes that display both variation in DNA methylation (within promoter region) and gene expression. (**A**) 153 genes with changes in granulocytes were used as input for the interaction analyses (see Material and Methods). Edges represent known or predicted protein-protein interactions, e.g. from curated databases and experiments. (**B**) Selected hub protein encoded by the *CANX* gene (calnexin, calcium-binding protein), which has 12 known interactions with other products of DMG/DEG genes from granulocytes. (**C**) Selected hub protein encoded by the CEBP gene (CCAAT Enhancer Binding Protein Beta), which has 11 known interactions with other products of DMG/DEG genes from granulocytes.

### Regulating the regulators – LOY-associated effect on transcription factor binding

We used the promoter sequences of the identified DMG/DEG genes to determine if they possess any enriched transcription factor binding sites (TFBS) as compared to genes without significant change in DNA methylation between LOY and ROY groups (see Methods). In granulocytes, we found 190 enriched motifs (153 among hypomethylated genes, 37 for hypermethylated ones; Supplementary File 1, Table S12). In monocytes, 72 motifs were enriched among promoters of the studied genes (59 for hypomethylated and 13 for hypermethylated genes; Supplementary File 1, Table S13).

Among the top-ranking motifs in hypomethylated genes, both in granulocytes and monocytes, were those belonging to the SP transcription factors (SP1, SP2, SP3, and SP4). In granulocytes, the SP2 motif was ∼1.6 times more frequent in promoters of hypomethylated DMG/DEG genes, compared to the background set of genes (FDR 2.91e-165). Interestingly, SP2 transcription factor itself was identified among the genes with a significant change in methylation and expression in granulocytes (hypomethylated, upregulated). Another interesting example was the E2F4 transcription factor, whose motif was enriched among hypermethylated genes in granulocytes.

## DISCUSSION

Knowledge accumulated over the past decade on LOY suggests a causative effect of this aneuploidy on the pathogenesis of AD. Age is also the key factor associated with both LOAD and LOY in men. Despite that the two phenotypes are age-dependent, and the fact that LOY is strongly and independently associated with LOAD, the molecular mechanisms underlying the role of LOY in LOAD are not well studied. Furthermore, little is known about the potentially LOY-induced methylation effects that may underlie the observed LOY-associated transcriptome changes [4]. For that reason, we have studied sorted subsets of leukocytes derived from male LOAD patients for changes of CpG-methylation and corresponding effects on the gene expression. As the microglia in brain and monocytes in blood circulation represent functionally related cells [27–30, 66], the rationale was to focus on myeloid cell lineage from AD patients to elucidate the role of LOY in AD. Moreover, granulocytes and monocytes have been shown to be affected by LOY in AD patients as shown in our previous study [4]. The samples studied here contained either high levels of mosaic LOY (>=30% of cells) or were predominantly not affected by LOY, thus forming two groups used for comparisons. Moreover, we have related the changes in the CpG methylation with RNA analysis, which were performed in the same samples for many patients, allowing identification of up- and down-regulated genes, presumably as a consequence of change in the methylation state.

Granulocytes and monocytes from AD patients showed a similar percentage of LOY cells (Fig. 2A), which is consistent with frequent clonal expansions of LOY-cells in the myeloid lineage [4, 7]. By analyzing DNA methylation patterns in 39 granulocyte and 24 monocyte samples, we show herein a predominant hypomethylation in LOY cells in both studied subpopulations of cells. The overall number of significant differentially methylated genes (DMGs) was higher in granulocytes than in monocytes (15269 vs 340 DMGs, Fig 3A). Noteworthy is that the majority of hypomethylated genes in monocytes (231 out of 309) were shared with hypomethylated genes found in granulocytes. This implies that the presumed LOY-induced methylation effects could have a core of defined targets regardless of the cell type. In contrast to predominant hypomethylation, gene expression analysis showed that the ratio between of up and down regulated genes is comparable in granulocytes and monocytes (Fig. 4A). Interestingly, 1096 significant differentially expressed genes were shared between granulocytes and monocytes, 475 were downregulated and 621 were upregulated (Supplementary File 1, tables S4 and S5). This large overlap of genes with the same direction of change in expression supports hypothesis that LOY could have gene-specific impact on transcriptome in myeloid and perhaps other types of cells. An important finding from the combined analyses of methylation and gene expression is the dysregulation of DEGs, presumably caused by methylation changes. We identified 643 genes in granulocytes and 37 genes in monocytes, which had significantly altered methylation and expression patterns (Fig. 4B). According to the canonical model, hypomethylation within CpG islands and promoters is associated with increased expression, whereas hypermethylation results in diminished expression of the corresponding gene [39, 40]. In the comparison of LOY vs. ROY cells, we identified numerous clear cases of DMG and DEG pairs fitting the canonical model (Figure 5, Supplementary File 1, Tables S8). An interesting candidate gene linking differential methylation, LOY and male health is the *RARA* gene, which is hypomethylated and upregulated in LOY cells. The *RARA* gene, encoding retinoic acid receptor alpha (RARα) that binds retinoic acid (RA), activates a signaling cascade leading to the transition of promyelocytes into the mature white blood cells through the granulocytic line [67, 68]. Low levels of ligand result in upregulation of *RARA* expression and consequently lead to inhibition of the differentiation of mature myeloid cells. High expression of *RARA* can also influence leukocyte differentiation through its interaction with C/EBPβ [69]. C/EBPβ is an essential transcription factor during emergency granulopoiesis, an immunological response induced by pathogens, including viral disease [60]. It should also be mentioned that independent GWAS study identified SNP variant in the *C/EBPβ* gene is among the loci that predispose to LOY [2]. Moreover, it was shown that posttranslational modifications of C/EBP-β can influence lymphoid to myeloid cell trans-differentiation [70]. Our finding that both genes important for leukocyte differentiation (*RARA* and *C/EBPβ*) are hypomethylated and upregulated in LOY cells implies that LOY might significantly impact on the maturation of myeloid lineage. Intriguingly, our independent studies have revealed that LOY appears abundantly in cells of the first line of immune defense, i.e. the innate immune response [4, 7]. These cells are mainly monocytes and neutrophils, with the latter constituting the largest population of granulocytes and having the second-highest rate of daily cell turnover [71]. Thus, LOY may drive selective maturation pathways in myeloid cells through epigenetic mechanisms regulating key differentiation regulators like *RARA* and *CEBPB*. Another example of a myeloid differentiation factor gene found to be hypomethylated and upregulated in our analysis is *MYADM*, which exhibits augmented expression during myeloid cell formation [65, 72]. Therefore, our results might be converted into a marker for immature hematopoietic cells committed toward myelopoiesis.

Our analysis also revealed that granulocytes with LOY show higher expression of *FLI1* and *RUNX1*. Both genes are differentially methylated in LOY cells with *FLI1* being hypomethylated and *RUNX1* hypermethylated. FLI1 is a transcription factor, whose higher expression favors the differentiation into megakaryocytes rather than erythroid cells [73]. FLI1 interacts with RUNX1 and GATA2, which play a key role in the development of the hematopoietic system. Notably, *RUNX1* expression is following the upregulation of *FLI1* [73]. Independent genome-wide association studies have shown that regions enriched in LOY heritability overlapped with binding sites of various transcription factors, among them FLI1, RUNX1, and GATA2 [73]. In our study, we detected LOY-related hypomethylation of the NF-E2 gene, which is another transcription factor functioning within the FLI1 network [74]. It has been shown that the triad of transcriptional factors, NF-E2, FLI1, and RUNX1 cooperate in regions of dynamic chromatin in the late-differentiation stage of megakaryocytes in a mouse model [75]. Excessive expression of NF-E2 in CD34+ causes the expansion of erythroid progenitor cells, a consequence of delays in the early phase of erythroid maturation [76]. Furthermore, overexpression of *NF-E2* in a murine myeloproliferative neoplasm model causes several hematopoietic phenotypes, such as leukocytosis and excessive thrombocytosis, along with a chronic inflammation creating an oxidative stress environment [77]. Notably, increased production of reactive oxygen species is related to excessive activation of neutrophils, which occurs in AD patients [33, 78]. It has been observed that high levels of reactive oxygen species (ROS) lead to rapid accumulation of 8-oxoG in DNA, which triggers cellular responses and among them, increased expression of *OGG1* [79]. This factor regulates the expression of inflammatory cytokine-encoding genes such as CCL20, IL-1B, TNFa, CXCL1, or CXCL2 [80, 81]. The molecular mechanism is based on the binding of OGG1 to 8-oxoGs located in inflammation-related genes facilitating the binding of NFkB transcription factor and their expression [82]. Considering the above, the hypomethylation of the *OGG1* gene detected in our analysis, manifested by its increased expression could lead to an exacerbated inflammatory response of the immune system.

Another noteworthy aspect is that leukocytes with LOY derived from AD patients may possibly exhibit apoptosis-related processes. Indeed, our analysis revealed that genes involved in the regulation of apoptosis, in particular *TRADD*, *TNFRSF1A*, and *DFFB*, were differentially methylated. TRADD, the TNF receptor-related death domain, through interactions with the intracellular death domain (DD), binds to activated TNFRSF1A (tumor necrosis factor receptor superfamily member 1A), leading to programmed death and NF-κB activation [83]. TNFRSF1A, expressed in neutrophils, plays a key role in TNFα-adapted apoptosis, and its blocking has anti-apoptotic effects [84]. It has been observed that proapoptotic TRADD was upregulated at the transition from myelocytes/metamyelocytes (MYs) to mature neutrophils in humans [85]. Additionally, DFFB which codes caspase-dependent DNase, triggers apoptosis through DNA fragmentation and chromatin condensation.

LOY at a single-cell level is a binary event removing the entire chromosome, resulting in loss of ∼2% of the haploid genome [86], which likely influence the packaging of DNA and chromosomes within a nucleus. Specifically, *KDM5D* and *UTY* (also called *KDM6C*) have histone demethylase activity and loss of these functions due to LOY could have a profound impact on epigenetic regulation in leukocytes. For instance, KDM5D targets H3K4me3 of histones [87] and this chromatin landmark is usually found near TSSs, which is an indicator of transcriptionally active genes. Thus, in the event of LOY, we could expect changes at the level of gene expression, triggered by chromatin remodeling. Recent studies showed dysregulation of epigenetic processes in AD and that regulation may occur through chromatin higher-order structures [48, 49, 88].

In conclusion, we provide further evidence suggesting that LOY in immune cells plays a role in the pathogenesis of LOAD in men. Our combined analyses of CpG methylation as well as expression analysis identified new candidate genes and we confirm numerous genes already implicated in the pathogenesis of LOAD. The results are also well aligned with the hypothesis that age-related dysfunction of the immune system cells is one of the major factors contributing to the development of AD. LOY is also reflected in higher-level epigenetic changes that show an AD-specific pattern, further contributing as a potential biomarker of the disease.

## Abbreviations

LOY: loss of chromosome Y

ROY: retention of chromosome Y

AD: Alzheimer’s disease

DMP: differentially methylated probe

DEG: differentially expressed gene

DMG: differentially methylated gene

TSS: transcription start site

LOAD: late onset Alzheimer’s disease

EOAD: early onset Alzheimer’s disease

RNA-seq bulk: RNA sequencing

PBMCs: peripheral blood mononuclear cells

mLRRY: median Log R Ratio values of probes from the male-specific part of chromosome Y

FACS: fluorescence-activated cell sorting.

## Data Availability

The DNA methylation data used in this study are available from the authors upon a reasonable request. The bulk RNA-seq datasets are available upon a reasonable request from the authors of the original publication (PMID: 33837451).

## AUTHOR CONTRIBUTIONS

Conceptualization: J.P.D.; Resources & sample collection: E.R-B., H.D., D.S., M.S., J.B., K.W., J.J., J.R., V.G., N.F., A.K-R., L.K., M.I., J.P.D.; Methodology & experiments: M.J., B.B-O., E.R-B., H.D., D.S., M.S., J.B., K.W., J.J., J.R., V.G., N.F., A.K-R., L.K., M.I., J.P.D.; Data Analysis: M.J., B.B-O., J.P.D.; D.S., E.R-B.; Writing-Original Draft: M.J., B.B-O., J.P.D.; Writing-Review & Editing: all co-authors; Supervision: M.J., N.F., J.P.D.; Project Administration: N.F., J.P.D.; Funding Acquisition: J.P.D.

## CODE AVAILABILITY

Unless otherwise stated, all the data processing and visualization of the results was done in R (version 3.6.3). The code used is available from https://github.com/jakalssj3/LOY-AD-meth

## ETHICS APPROVAL

The study was conducted according to the guidelines of the Declaration of Helsinki. The study was approved by the local research ethics committee in Uppsala, Sweden (Regionala Etikprövningsnämnden in Uppsala (EPN): Dnr 2005-244, Ö48-2005; Dnr 2013/350; Dnr 2015/092; Dnr 2015/458; Dnr 2015/458/2, the latter with update from 2018) and the Bioethical Committee of the Regional Medical Chamber in Kraków, Poland (No. 6/KBL/OIL/2014).

## CONSENT TO PARTICIPATE

All participants or next of kin have given their written informed consent to participate.

### ACKNOWLEDGEMENTS

We thank the patients and healthy controls for sample contribution and information provided in the questionnaire. We are grateful to Dr. Jakub Mieczkowski for the project guidance and Dr. Eva Tiensuu Janson for critical review of the manuscript and Dr. Lars A Forsberg and Dr. Jonatan Halvardson for access to RNA-seq data.

## SUPPLEMENTARY FIGURES

**Figure S1.**
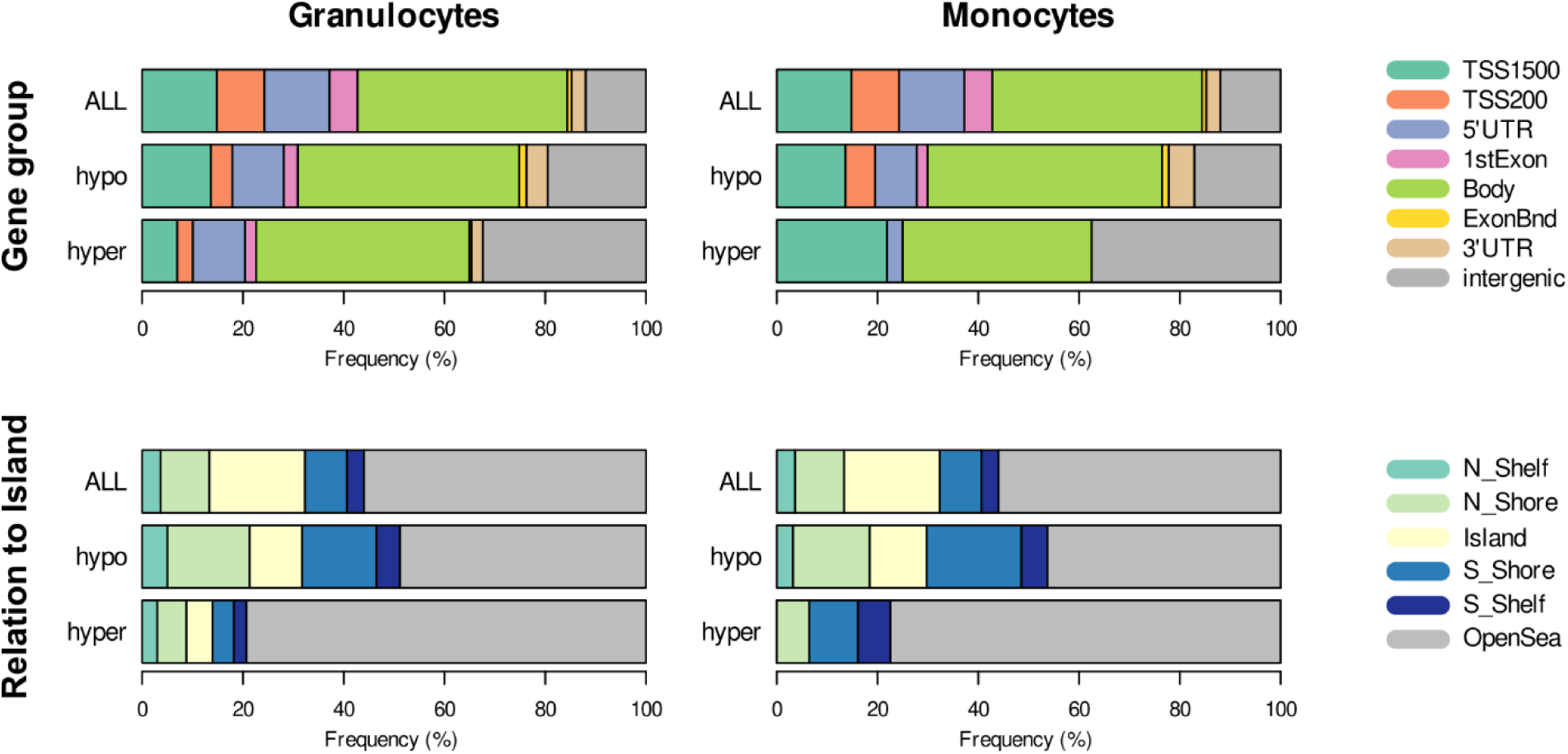
Genomic distribution of the identified hyper- and hypomethylated DMPs as compared to the distribution of all probes on the EPIC chip. **Top panel.** Distribution of probes and DMPs in relation to gene annotations (UCSC_RefGene_Group column in the Infinium MethylationEPIC Manifest). The left and right panels correspond to granulocytes and monocytes, respectively. **Bottom panel.** Distribution of probes and DMPs in relation to CpG islands (Relation_to_Island column in the Infinium MethylationEPIC Manifest).

**Figure S2.**
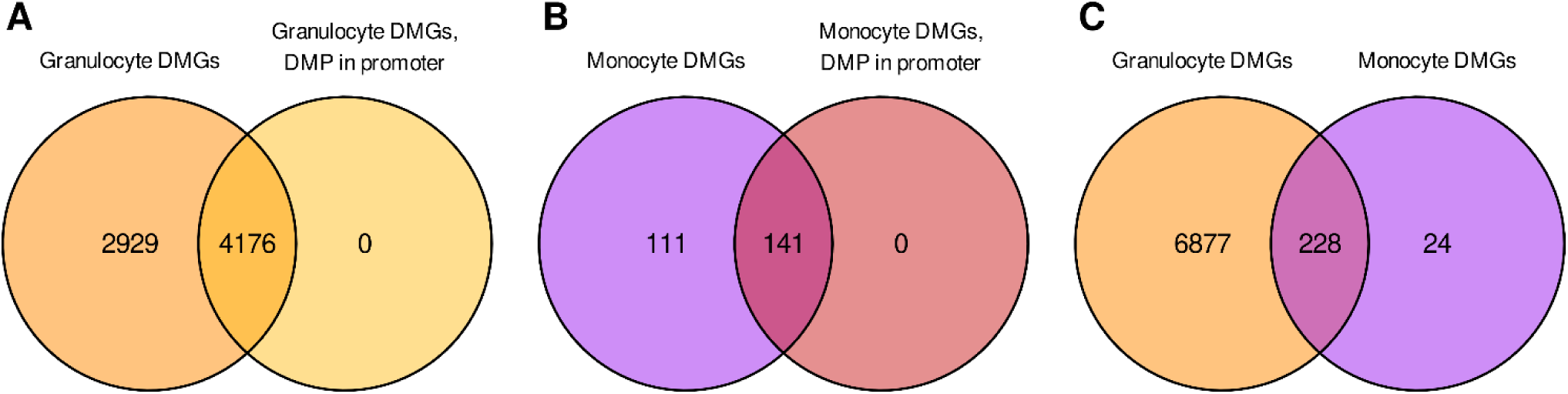
Intersections of gene sets identified to have differentially methylated probes (DMPs) identified in them (hence called DMGs). (**A**) Intersection of DMGs found in Granulocytes. (**B**) Intersection of DMGs found in Monocytes. (**C**) Intersection of DMGs identified in Granulocytes and Monocytes.

**Figure S3.**
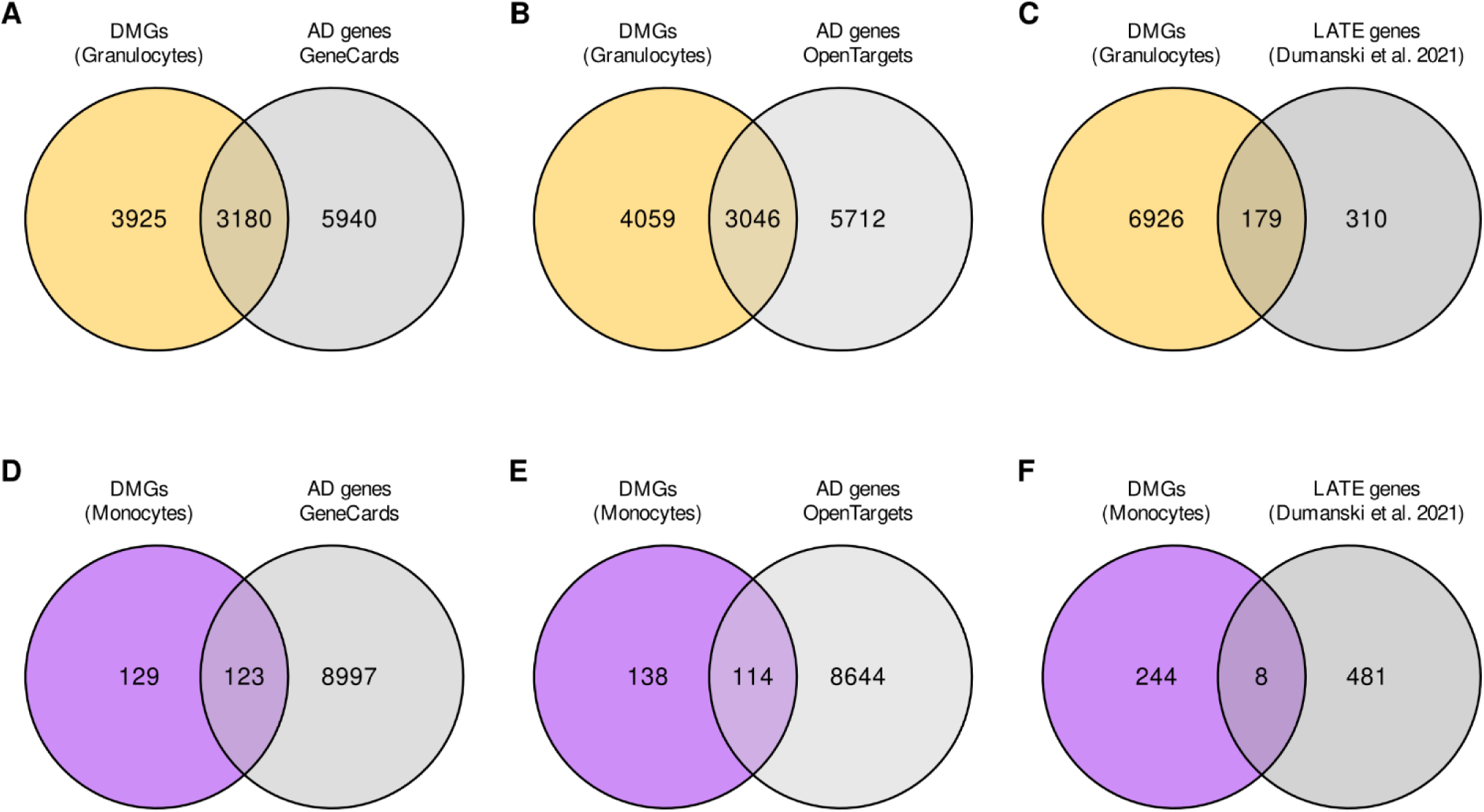
Intersection of genes containing differentially methylated probes (DMGs) with sets of genes identified as related to Alzheimer’s disease. (**A**) Intersection of granulocyte DMGs with AD related genes from GeneCards database (**B**) Intersection of granulocyte DMGs with AD related genes from OpenTargets database (**C**) Intersection of granulocyte DMGs with so-called LATE genes from Dumanski et. al 2021 (**D**) Intersection of monocyte DMGs with AD related genes from GeneCards database (**E**) Intersection of monocyte DMGs with AD related genes from OpenTargets database (**F**) Intersection of monocyte DMGs with LATE genes.

